# Epidemiology and Transmission of COVID-19 in Shenzhen China: Analysis of 391 cases and 1,286 of their close contacts

**DOI:** 10.1101/2020.03.03.20028423

**Authors:** Qifang Bi, Yongsheng Wu, Shujiang Mei, Chenfei Ye, Xuan Zou, Zhen Zhang, Xiaojian Liu, Lan Wei, Shaun A. Truelove, Tong Zhang, Wei Gao, Cong Cheng, Xiujuan Tang, Xiaoliang Wu, Yu Wu, Binbin Sun, Suli Huang, Yu Sun, Juncen Zhang, Ting Ma, Justin Lessler, Tiejian Feng

**Affiliations:** Johns Hopkins Bloomberg School of Public Health, Baltimore, United States; Shenzhen Center for Disease Control and Prevention, Shenzhen, China; Harbin Institute of Technology at Shenzhen, Shenzhen, China; Peng Cheng Laboratory, Shenzhen, China

## Abstract

**Background:** Rapid spread of SARS-CoV-2 in Wuhan prompted heightened surveillance in Shenzhen and elsewhere in China. The resulting data provide a rare opportunity to measure key metrics of disease course, transmission, and the impact of control.

**Methods:** The Shenzhen CDC identified 391 SARS-CoV-2 cases from January 14 to February 12, 2020 and 1286 close contacts. We compare cases identified through symptomatic surveillance and contact tracing, and estimate the time from symptom onset to confirmation, isolation, and hospitalization. We estimate metrics of disease transmission and analyze factors influencing transmission risk.

**Findings:** Cases were older than the general population (mean age 45) and balanced between males (187) and females (204). Ninety-one percent had mild or moderate clinical severity at initial assessment. Three have died, 225 have recovered (median time to recovery is 21 days). Cases were isolated on average 4.6 days after developing symptoms; contact tracing reduced this by 1.9 days. Household contacts and those travelling with a case where at higher risk of infection (ORs 6 and 7) than other close contacts. The household secondary attack rate was 15%, and children were as likely to be infected as adults. The observed reproductive number was 0.4, with a mean serial interval of 6.3 days.

**Interpretation:** Our data on cases as well as their infected and uninfected close contacts provide key insights into SARS-CoV-2 epidemiology. This work shows that heightened surveillance and isolation, particularly contact tracing, reduces the time cases are infectious in the community, thereby reducing *R*. Its overall impact, however, is uncertain and highly dependent on the number of asymptomatic cases. We further show that children are at similar risk of infection as the general population, though less likely to have severe symptoms; hence should be considered in analyses of transmission and control.

## Introduction

Since emerging in Wuhan, China in December of 2019^1^, the epidemic of the novel coronavirus SARS-CoV-2 has progressed rapidly. The disease caused by this virus, dubbed COVID-19 (coronavirus disease 2019) by the World Health Organization (WHO) is characterized by fever, cough, fatigue, shortness of breath, pneumonia, and other respiratory tract symptoms ^2–4^, and in many cases progresses to death. As of February 24, 2020, there have been 79,331 confirmed cases and 2,618 deaths reported worldwide^5^. The vast majority of these remain confined to Hubei province, but there has been significant spread elsewhere in China and the world. A rapid and robust response by the global scientific community has described many key aspects of SARS-CoV-2 ^1,2,6–8^ transmission and natural history, but key questions remain.

If well tracked, early introductions of an emerging pathogen provide a unique opportunity to characterize its transmission, natural history, and the effectiveness of screening. The careful monitoring of cases and low probability of infection from the general community enables inferences, critical to modeling the course of the outbreak, that are difficult to make during a widely disseminated epidemic. In particular, we are able to make assumptions about when and where cases were likely infected that are impossible when the pathogen is widespread. Furthermore, during these early phases, uninfected and asymptomatic contacts are often closely tracked, providing critical information on transmission and natural history. Combined, this data on early introductions can be used to give insights into disease natural history ^9^, transmission characteristics ^10^, and the unseen burden of infection ^11^.

Here, we use data collected by the Shenzhen Center for Disease Control and Prevention (Shenzhen CDC) on 391 cases of COVID-19 and 1,286 of their close contacts to characterize key aspects of its epidemiology outside of Hubei province. We characterize differences in demographics and severity between cases identified through symptom-based surveillance and the monitoring of close case contacts, and estimate the time to key events, such as confirmation, isolation, and recovery. Using data from contact tracing, we characterize SARS-CoV-2 transmission by estimating key values, such as the household secondary attack rate, serial interval and observed reproductive number.

## Methods

### Case Identification

On January 8, 2020, Shenzhen CDC identified the first case of pneumonia with unknown cause and began monitoring travelers from Hubei province for symptoms of COVID-19. Over the next two weeks this surveillance program expanded to include travelers from Hubei regardless of symptoms, patients at local hospitals, and individuals detected by fever screening in neighborhoods and at local clinics. Suspected cases and close contacts were tested for SARS-CoV-2 by PCR of nasal swabs at 28 qualified local hospitals, 10 district level CDCs, and 2 third party testing organizations, with final confirmation performed at the Guangdong CDC or Shenzhen CDC (Text S1). Close contacts were defined as those who lived in the same apartment, shared a meal, traveled, or socially interacted with an index case during the period starting two days before symptom onset. Casual contacts (e.g., other clinic patients) and some close contacts (e.g., nurses) who wore a mask during exposure were not included in this group.

Symptomatic cases were isolated and treated at designated hospitals regardless of test results. Asymptomatic positives were isolated at centralized facilities. Close contacts and travelers from Hubei testing negative were isolated at home or a central facility, and monitored for 14 days. PCR testing was required for all close contacts at the beginning of isolation, and release was conditional on a negative PCR result. Basic demographics, signs and symptoms, clinical severity, and exposure history were recorded for all confirmed cases.

Here, we analyze confirmed cases identified by the Shenzhen CDC between Jan 14, 2020 and Feb 12, 2020, and the close contacts of cases confirmed before February 9th.

### Epidemiologic and Clinical Characteristics of Cases

We define symptom-based surveillance to include symptomatic screening at airport and train stations, community fever monitoring, home observation of recent travellers to Hubei, and testing of hospital patients. Contact-based surveillance is the identification of cases through monitoring and testing of close contacts of confirmed cases. By protocol, those in the contact-based group were tested for SARS-CoV-2 infection regardless of symptoms, while those in the other categories were tested only if they showed signs or symptoms of disease.

At first clinical assessment, data were recorded on 21 signs and symptoms (see supplement), and disease severity was assessed. Cases with fever, respiratory symptoms, and radiographic evidence of pneumonia were classified as having *moderate* symptoms. Cases were classified as having *severe* symptoms if they had any of: breathing rate ≥30/min; oxygen saturation level ≤93% at rest; oxygen concentration level PaO_2_/FiO_2_ ≤ 300mmHg (1mmHg=0.133kPa); lung infiltrates >50% within 24-48 hours; respiratory failure requiring mechanical ventilation; septic shock; or multiple organ dysfunction/failure. All other symptomatic cases were classified as *mild*.

Relationships between demographics, mode of detection and symptom severity were assessed and characterized using χ^2^-tests, and logistic regression.

### Timing of Key Events

Distributions were fit to the timing of key events in each confirmed case’s course of infection and treatment. The time from infection to symptom onset (incubation periods) were assumed to be log-normally distributed and estimated as previously described ^12–14^. Cases who recently travelled to Hubei were assumed to have been exposed while there. Cases without a recent travel history but with exposure to a confirmed case, were assumed to be exposed from the time of earliest to latest possible contact with that case. Only cases for whom we could bound the earliest and latest period of exposure and had a date of symptom onset were included in the analysis.

Time between symptom onset and recovery was estimated using parametric survival methods. Patients who had not recovered were considered to be censored on February 22, 2020 or at the time of death. All other delay distributions were estimated by directly fitting parametric distributions to time between symptom onset or arrival in Shenzhen, and confirmation, isolation or hospitalization. Confidence intervals were calculated using bootstrapping or standard parametric estimators ^15^.

### Transmission Characteristics

Transmission was characterized by examining the relationship between confirmed cases and their infected and uninfected close contacts. The household secondary attack rate (SAR) was calculated as the percentage of household contacts (those sharing a room, apartment or other sleeping arrangement) who were later confirmed to be infected with SARS-CoV-2. The distribution of serial intervals (the time between symptom onset in infector and infectee) was calculated by fitting parametric distributions to the time of symptom onset in clear infector/infectee pairs. The mean observed reproductive number, *R*, and distribution of personal reproductive numbers (i.e., the number of secondary infections caused by each case) were calculated from the number of secondary infections observed among close contacts of each index case, with ambiguities resolved through multiple imputation. The relative odds of transmission among contacts of various types were estimated using conditional logistic regression and random effects models, to account for differing numbers of possible infectors in each risk group. Confidence intervals were estimated using bootstrapping or standard parametric approaches.

### Ethics Statement

This work was conducted in support of an ongoing public health response, hence was determined not to be human subjects research after consultation with the Johns Hopkins Bloomberg School of Public Health IRB. Analytic datasets were constructed in an anonymized fashion, and all analysis of personally identifiable data took place on site at the Shenzhen CDC.

## Results

### Epidemiologic and Clinical Characteristics

Between Jan 14, 2020 and Feb 12, 2020 the Shenzhen CDC confirmed 391 cases of SARS-CoV-2 infection (Table 1). Of 379 with a known mode of detection, 77% were detected through symptom-based surveillance. Overall, there were approximately equal numbers of male and female cases (187 vs. 204), and 79% were adults between the ages of 30 and 69. At the time of first clinical assessment, most cases were mild (26%) or moderate (65%); and only 35 (9%) were severe. Eighty-four percent of cases had fever at the time of initial assessment, while 6% had no signs or symptoms. As of February 22, 2020, final clinical outcomes were known for 228 of the 391 cases in our data; with three having died and 225 recovered.

**Table 1:** Demographic and clinical characteristics of cases by contact-based vs. symptom-based surveillance.

| Variable | Value | Contact-based surveillance (N=87) | Symptom-based surveillance (N=292) | Unknown/other (N=12) | Total (N=391) | P-value |
| --- | --- | --- | --- | --- | --- | --- |
| sex | female | 63 (72.4%) | 131 (44.9%) | 10 (83.3%) | 204 (52.2%) | <0.001 |
|  | male | 24 (27.6%) | 161 (55.1%) | 2 (16.7%) | 187 (47.8%) |  |
| age | 0-9 | 13 (14.9%) | 6 (2.1%) | 1 (8.3%) | 20 (5.1%) | <0.001 |
|  | 10-19 | 5 (5.7%) | 6 (2.1%) | 1 (8.3%) | 12 (3.1%) |  |
|  | 20-29 | 11 (12.6%) | 23 (7.9%) | 0 (0.0%) | 34 (8.7%) |  |
|  | 30-39 | 15 (17.2%) | 71 (24.3%) | 1 (8.3%) | 87 (22.3%) |  |
|  | 40-49 | 9 (10.3%) | 49 (16.8%) | 2 (16.7%) | 60 (15.3%) |  |
|  | 50-59 | 10 (11.5%) | 63 (21.6%) | 1 (8.3%) | 74 (18.9%) |  |
|  | 60-69 | 20 (23.0%) | 60 (20.5%) | 6 (50.0%) | 86 (22.0%) |  |
|  | 70+ | 4 (4.6%) | 14 (4.8%) | 0 (0.0%) | 18 (4.6%) |  |
| severity | mild | 18 (20.7%) | 82 (28.1%) | 2 (16.7%) | 102 (26.1%) | 0.03 |
|  | moderate | 66 (75.9%) | 180 (61.6%) | 8 (66.7%) | 254 (65.0%) |  |
|  | severe | 3 (3.4%) | 30 (10.3%) | 2 (16.7%) | 35 (9.0%) |  |
| symptomatic | no | 17 (19.5%) | 8 (2.7%) | 0 (0.0%) | 25 (6.4%) | <0.001 |
|  | yes | 70 (80.5%) | 284 (97.3%) | 12 (100.0%) | 366 (93.6%) |  |
| fever | no | 25 (28.7%) | 34 (11.6%) | 2 (16.7%) | 61 (15.6%) | <0.001 |
|  | yes | 62 (71.3%) | 258 (88.4%) | 10 (83.3%) | 330 (84.4%) |  |

Cases detected through symptom-based surveillance were more often male (55% vs 28%) and between the ages of 20-69 (91% vs 75%) than those detected through contact-based surveillance (Tables 1 and 2). At the time of first clinical assessment, 29% of the contact-based surveillance group did not have fever, and 20% had no symptoms. In contrast, 88% of the symptom-based surveillance group had fever, and only 8 reported no symptoms.

**Table 2:** Association of clinical and demographic factors with mode of detection.

|  |  | Outcome: symptom-based surveillance |  |  |  |  |  | Outcome: moderate/severe symptom |  |  |  |  |  |
| --- | --- | --- | --- | --- | --- | --- | --- | --- | --- | --- | --- | --- | --- |
|  |  | Univariate regression |  |  | Multivariate regression |  |  | Univariate regression |  |  | Multivariate regression |  |  |
|  |  | OR | 2.5% | 97.5% | OR | 2.5% | 97.5% | OR | 2.5% | 97.5% | OR | 2.5% | 97.5% |
| sex | female | ref |  |  | ref |  |  | ref |  |  | ref |  |  |
|  | male | 3.23 | 1.93 | 5.53 | 3.06 | 1.77 | 5.44 | 1.30 | 0.82 | 2.07 | 1.37 | 0.85 | 2.22 |
| age | 0-9 | 0.07 | 0.02 | 0.23 | 0.08 | 0.02 | 0.25 | 0.88 | 0.30 | 2.77 | 0.67 | 0.22 | 2.23 |
|  | 10-19 | 0.19 | 0.05 | 0.77 | 0.17 | 0.04 | 0.76 | 1.08 | 0.28 | 5.27 | 0.88 | 0.22 | 4.40 |
|  | 20-29 | 0.33 | 0.12 | 0.89 | 0.37 | 0.13 | 1.02 | 0.65 | 0.28 | 1.56 | 0.61 | 0.26 | 1.49 |
|  | 30-39 | 0.75 | 0.31 | 1.77 | 0.8 | 0.32 | 1.93 | 1.17 | 0.58 | 2.37 | 1.15 | 0.57 | 2.33 |
|  | 40-49 | 0.86 | 0.32 | 2.33 | 0.79 | 0.29 | 2.19 | 1.27 | 0.58 | 2.83 | 1.21 | 0.55 | 2.72 |
|  | 50-59 | ref |  |  | ref |  |  | ref |  |  | ref |  |  |
|  | 60-69 | 0.48 | 0.2 | 1.08 | 0.44 | 0.18 | 1.04 | 1.50 | 0.72 | 3.16 | 1.43 | 0.68 | 3.03 |
|  | 70+ | 0.56 | 0.16 | 2.25 | 0.45 | 0.12 | 1.89 | 1.41 | 0.45 | 5.43 | 1.28 | 0.40 | 4.96 |
| severity | mild | ref |  |  | ref |  |  | - | - | - | - | - | - |
|  | moderate | 0.6 | 0.33 | 1.05 | 0.51 | 0.26 | 0.94 | - | - | - | - | - | - |
|  | severe | 2.2 | 0.68 | 9.84 | 1.51 | 0.42 | 7.23 | - | - | - | - | - | - |
| fever | no | ref |  |  | - | - | - | ref |  |  | - | - | - |
|  | yes | 3.06 | 1.69 | 5.49 | - | - | - | 0.94 | 0.49 | 1.75 | - | - | - |
| symptomatic | no | ref |  |  |  |  |  | ref |  |  |  |  |  |
|  | yes | 8.62 | 3.68 | 21.89 | - | - | - | 0.51 | 0.15 | 1.39 | 0.55 | 0.29 | 1.01 |
| surveillance method | contact-based | ref |  |  | - | - | - | ref |  |  | - | - | - |
|  | symptom-based | - | - | - | - | - | - | 0.67 | 0.37 | 1.17 | - | - | - |

In multiple logistic regression, severe symptoms were associated with being male (OR 2.5, 95% CI 1.1,6.1). There was a general trend of increasing probability of severe symptoms with age, though only 60-69 year olds showed a significant difference from the reference category (OR 3.4 versus 50-59 year olds; 95% 1.4,9.5) and being male (OR=2.5, 95%CI 1.1, 6.1) (Table 2).

### Timing of Key Events

Based on 183 cases with a well-defined period of exposure and symptom onset (Figure S1), we estimate the median incubation period for COVID-19 to be 4.8 days (95% CI 4.2,5.4) [Figure 2, Table S2], and that 95% of those who develop symptoms will do so within 14.0 days (95% CI 12.2,15.9) of infection.

Based on 228 cases with known outcomes, we estimate that the median time to recovery is 22 days (95% CI 21,24) in 50-59 year olds, and is estimated to be significantly shorter in younger adults (e.g., 19 days in 20-29 year olds), and longer in older groups (e.g., 23 days in those aged 60-69) despite not being statistically significant (Table S5, Figure S3). In multiple regression models including sex, age, baseline severity and method of detection, in addition to age, baseline severity was associated with time to recovery. Compared to those with mild symptoms, those with severe symptoms were associated with a 41% (95% CI, 24%,60%) increase in time to recovery. Thus far, only three have died. These occurred 35-44 days from symptom onset and 27-33 days from confirmation.

Cases detected through symptom-based surveillance were confirmed on average 5.5 days (95% CI 5.0, 5.9) after symptom onset (Figure 3, Table S2); compared to 3.2 days (95% CI 2.6,3.7) in those detected by contact-based surveillance. 17 cases were isolated before developing symptoms. Among those isolated after, the symptom-based surveillance group were, on average, isolated 4.6 days (95% CI 4.1,5.0) after symptom onset, versus 2.7 days (95% CI 2.1,3.3) in the contact-based surveillance group. Hence, contact-based surveillance was associated with a 2.3 days (95% CI 1.5,3.0) decrease in time to confirmation and a 1.9 days (95% CI 1.1,2.7) decrease in time to isolation. Timings between symptom onset and hospitalization were similar to isolation results (Figure 3, Table S2).

Sixty-four percent (191/298) of travelers developed symptoms after arriving in Shenzhen, with a mean time from arrival to symptom onset of 4.9 days (95% CI 4.2, 5.5) (Table S2). Those developing symptoms prior to arrival or on the day of arrival were confirmed as cases on average 4.5 days (95% CI 3.8,5.1) after arrival, and isolated on average 3.1 days (95% CI, 2.5,3.7) after.

### Transmission Characteristics

Overall, 1286 close contacts were identified for index cases testing positive for SARS-CoV-2 between January 14 and February 9, 2020, with 83% (244/292) of cases having at least one close contact. Ninety-five percent of close contacts were followed 12 days or longer. Ninety-eight tested PCR positive for SARS-CoV-2 infection, and one had presumptive infection. Excluding those with a missing test result, we found that the secondary attack rate was 15.8% (95% CI 12.9,19.4) among household contacts and 10.3% (95%CI 8.4,12.6) overall (these drop to 11.2% and 6.6% if those with missing results are considered to be negative). In multivariable analysis of contact types household contact (OR 6.3, 95% CI 1.5, 26.3) and travelling together (OR 7.1, 95% CI 1.4, 34.9) were significantly associated with infection. Reporting contact occurred “often” was also associated with increased risk of infection (OR 8.8 versus moderate frequency contacts, 95% CI 2.6,30.1).

Attack rates were similar across infectee age categories (Table 3), though there is some indication of elevated attack rates in older age groups (Figure 1). Notably, the rate of infection in children under 10 (7.4%) was similar to the population average (7.9%). There was no significant association between probability of infection and age of the index case. Surprisingly, in univariate analysis a longer time in the community prior to isolation was associated with a reduced risk of causing infections. However, this association was no longer significant after adjusting for contact frequency and type.

**Table 3:** Risk factors for SAR-CoV-2 infection among close contacts

|  |  |  |  |  | Univariate regression |  |  | Multivariate regression |  |  |
| --- | --- | --- | --- | --- | --- | --- | --- | --- | --- | --- |
|  |  | N | Infected | Attack rate %<br>(95% CI) | Odds ratio | 2.5% | 97.5% | Odds ratio | 2.5% | 97.5% |
| sex | female | 558 | 58 | 10.4 (8.1, 13.2) |  |  |  | - | - | - |
|  | male | 486 | 26 | 5.3 (3.7, 7.7) | 0.43 | 0.21 | 0.86 | - | - | - |
| age | 0-9 | 148 | 11 | 7.4 (4.2, 12.8) | 2.33 | 0.38 | 14.05 | - | - | - |
|  | 10-19 | 85 | 6 | 7.1 (3.3, 14.6) | 3.50 | 0.53 | 23.24 | - | - | - |
|  | 20-29 | 114 | 7 | 6.1 (3.0, 12.1) | 4.91 | 0.74 | 32.64 | - | - | - |
|  | 30-39 | 268 | 16 | 6.0 (3.7, 9.5) | 1.84 | 0.34 | 9.80 | - | - | - |
|  | 40-49 | 143 | 7 | 4.9 (2.4, 9.8) | 3.46 | 0.55 | 21.92 | - | - | - |
|  | 50-59 | 110 | 10 | 9.1 (5.0, 15.9) | ref |  |  | - | - | - |
|  | 60-69 | 130 | 20 | 15.4 (10.2, 22.6) | 5.68 | 1.01 | 32.09 | - | - | - |
|  | 70+ | 72 | 7 | 9.7 (4.8, 18.7) | 4.26 | 0.64 | 28.44 | - | - | - |
| HH | no | 456 | 4 | 0.9 (0.3, 2.2) | ref |  |  | ref |  |  |
|  | yes | 686 | 77 | 11.2 (9.1, 13.8) | 15.10 | 3.69 | 61.69 | 6.27 | 1.49 | 26.33 |
| travel | no | 824 | 63 | 7.6 (6.0, 9.7) | ref |  |  | ref |  |  |
|  | yes | 318 | 18 | 5.7 (3.6, 8.8) | 9.13 | 1.85 | 45.08 | 7.06 | 1.43 | 34.91 |
| meal | no | 435 | 20 | 4.6 (3.0, 7.0) | ref |  |  | ref |  |  |
|  | yes | 707 | 61 | 8.6 (6.8, 10.9) | 23.01 | 2.51 | 211.2 | 7.13 | 0.73 | 69.32 |
| contact frequency | rare | 230 | 1 | 0.4 (0.02, 2.4) | <0.0001 | 0 | Inf | - | - | - |
|  | moderate | 305 | 9 | 3.0 (1.6, 5.5) | ref |  |  | - | - | - |
|  | often | 555 | 71 | 12.8 (10.3, 15.8) | 8.8 | 2.58 | 30.06 | - | - | - |

**Figure 1:**
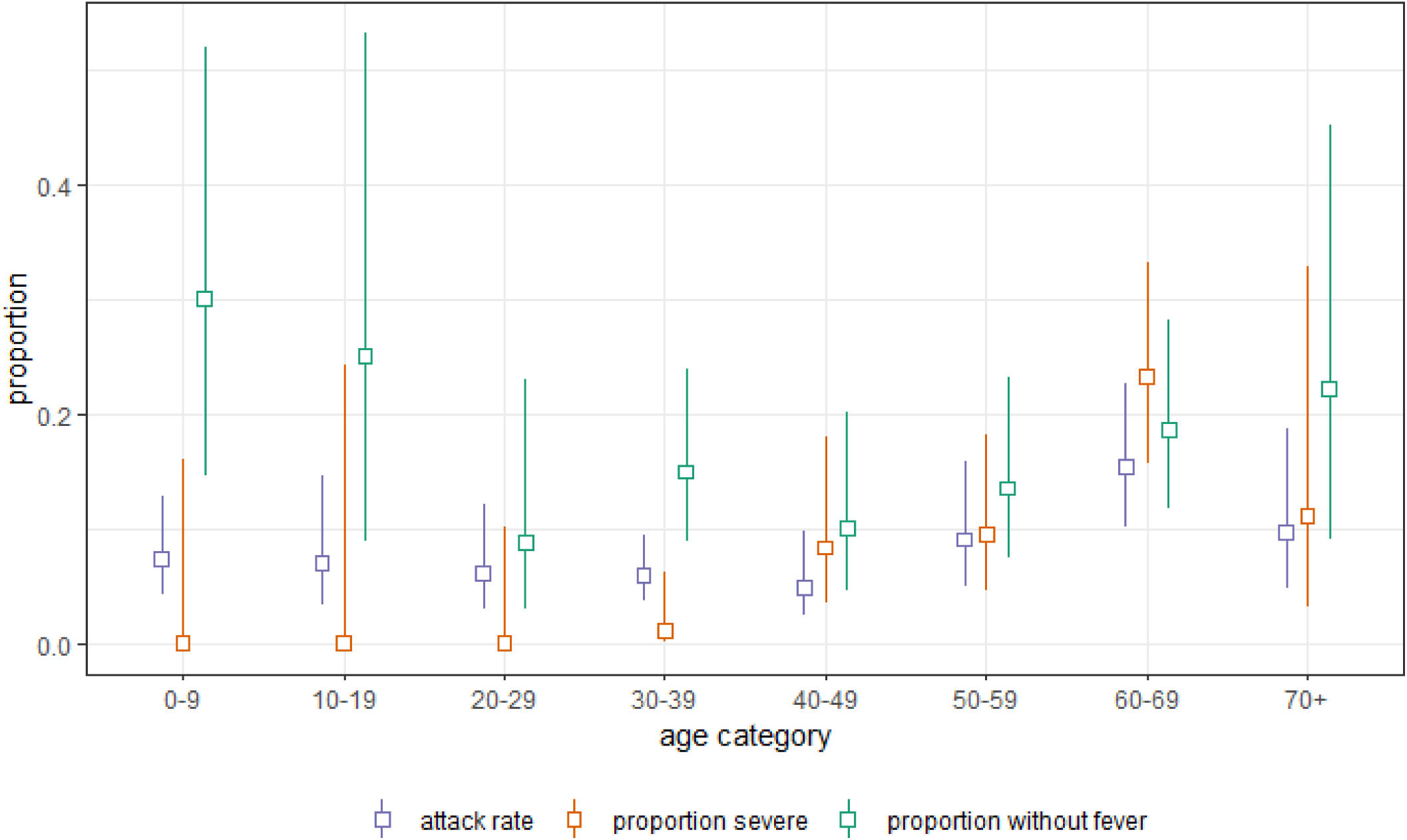
Attack rate among close contacts, baseline severity, and proportion without fever at initial assessment by age group.

Based on 48 pairs of cases with a clear infector-infectee relationship and time of symptom onset, we estimate that the serial interval is gamma distributed with mean 6.3 days (95% CI 5.2,7.6) and a standard deviation of 4.2 days (95% CI, 3.1,5.3) (Figure 2B, Table S2). Hence, 95% of cases are expected to develop symptoms within 14.3 (95% CI, 11.1,17.6) days of their infector. It should be noted this estimate includes the effect of isolation on truncating the serial interval. Stratified results show that if the infector was isolated less than 3 days after infection the average serial interval was 3.6 days, increasing to 8.1 days if the infector was isolated on the third day after symptom onset or later (Table S4).

The mean number of secondary cases caused by each index case (i.e., the observed reproductive number, *R*), was 0.4 (95% CI 0.3,0.5). The distribution of personal reproductive numbers was highly overdispersed, with 80% of infections being caused by 8.9% (95% CI 3.5,10.8) of cases (negative binomial dispersion parameter 0.58, 95% CI 0.35, 1.18).

**Figure 2:**
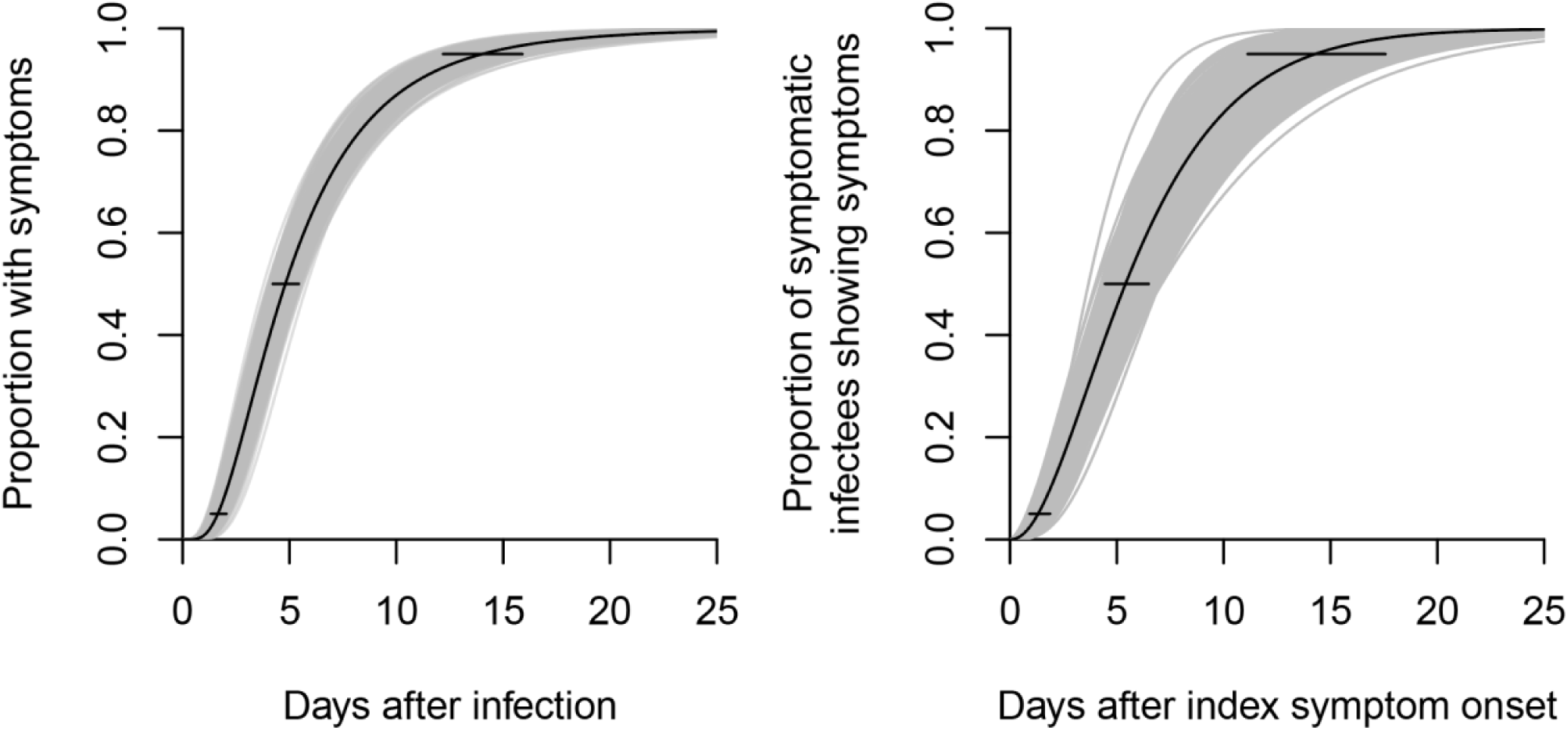
**(A) The proportion of cases having developed symptoms to COVID-19 by days after infection (i.e., the cumulative distribution function of the incubation period). (B) The proportion of cases infected by an index case who have developed symptoms by a given number of days after the day of symptom onset of the index case (i.e., the cumulative distribution function of the serial interval)**. The maximum-likelihood estimates for the parametric distribution of the cumulative distribution function are shown, along with 1000 parametric bootstrap estimates of the cumulative distribution function. We estimate the median incubation period of COVID-19 is 4.8 days (95%CI, 4.2, 5.4). 5% of the cases who develop symptoms will do so by 1.6 days (95% CI, 1.3, 2.0) after infection, and 95% by 14.0 days (12.2, 15.9). We estimated that the median serial interval of COVID-19 is 5.4 days (95% CI, 4.4 to 6.5). 5% of the infected who develop symptoms will do so by 1.3 days (95% CI, 0.9 to 1.9) after symptom onset of the index case, and in 95% by 14.3 days (95% CI, 11.1 to 17.6).

**Figure 3:**
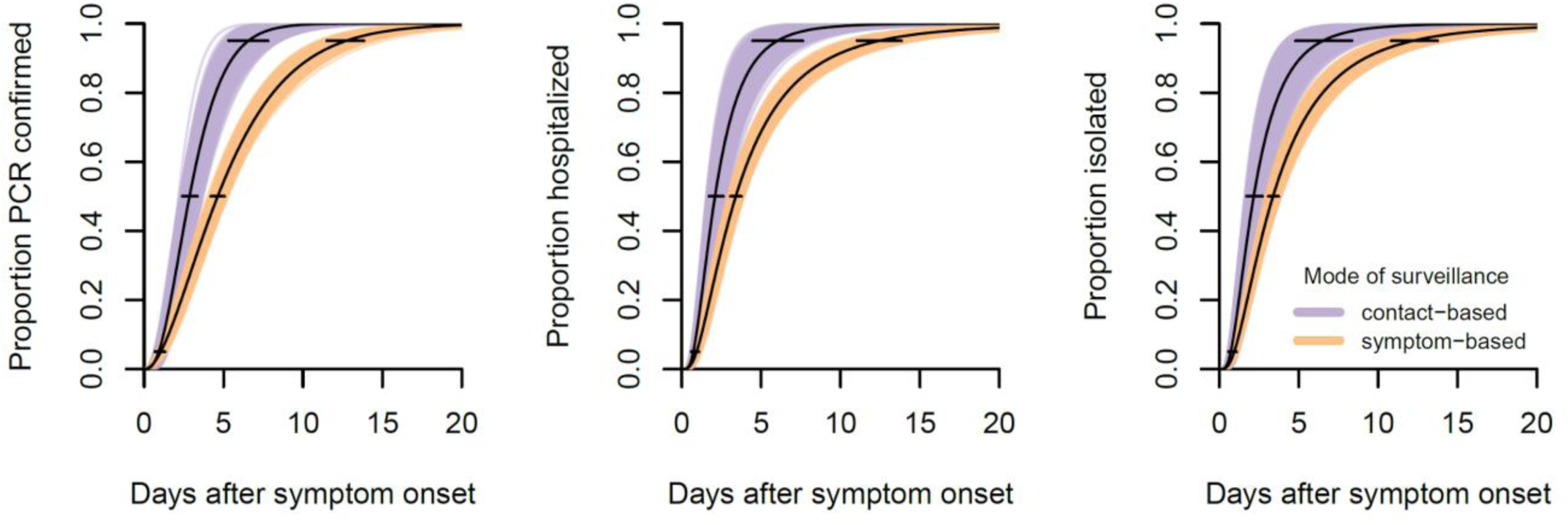
**Time between symptom onset and (A) SARS-CoV-2 confirmation, (B) hospitalization and (C) isolation among cases detected by contact-based and symptom-based surveillance**. The maximum-likelihood estimates for the parametric distribution of the cumulative distribution function are shown, along with 1000 parametric bootstrap estimates of the cumulative distribution function. **Panel A** shows estimates of the proportion of cases who are PCR confirmed, according to the number of days after symptom onset. We estimated that 50% of the cases detected through symptom-based surveillance were PCR confirmed by 4.6 days (95% CI, 4.2 to 5.0) after symptom onset, in 95% by 12.7 days (95% CI, 11.5 to 13.8) after symptom onset. Contact-based surveillance reduced the days from symptom onset to PCR confirmation to 2.9 days (95% CI, 2.4, 3.4) in 50% cases and 6.6 days (95%CI, 5.3, 8.0) in 95% cases. **Panel B** shows estimates of the proportion of cases who are hospitalized, according to the number of days after symptom onset. We estimated that 50% of the cases detected through symptom-based surveillance were hospitalized by 3.4 days (95% CI, 3.1 to 3.8) after symptom onset, in 95% by 12.4 days (95% CI, 10.9 to 13.8). Contact-based surveillance reduced the days from symptom onset to hospitalization to 2.1 days (95% CI, 1.7, 2.6) in 50% cases, and 6.0 days (95% CI, 4.5, 7.5) in 95% cases. **Panel C** shows estimates of the proportion of cases isolated, according to the number of days after symptom onset. We estimated that 50% of the cases detected through symptom-based surveillance were isolated by 3.4 days (95% CI, 3.1 to 3.7) after symptom onset, in 95% by 12.2 days (95% CI, 10.8 to 13.6). Contact-based surveillance reduced the days from symptom onset to isolation to 2.2 days (95% CI, 1.7, 2.6) in 50% cases, and 6.5 days (95% CI, 4.7, 8.2) in 95% cases.

### Potential Impact of Surveillance and Isolation on Transmission

To calculate the potential impact of surveillance and isolation on transmission, we considered a range of possible infectious periods where infectiousness varied over time. We define the mean infectious day (i.e., the average number of days after symptom onset an infector is expected to infect a secondary case) as the weighted mean of the infectious period, where each day is weighted by relative infectiousness. We consider periods where the mean infectious day is less than 15 days after symptom onset (roughly the period of SARS and early SARS-CoV-2 reports^16,17^), and assume that *R*=2.6 and that isolation effectively ends the infectious period. Under these assumptions we find if the mean infectious day is greater than 5 days, then it may be possible to bring *R* below one in those detected by symptom-based surveillance; and the same can be accomplished by contact-based surveillance if the mean infectious day is greater than 3 days. For the impact of passive surveillance alone to achieve our observed *R* of 0.4, we project the mean infectious day must be at least 5.5 days (and likely more) after symptom onset.

Even if transmission is completely eliminated in the group captured by surveillance (e.g., if we could get perfect surveillance on the day of symptom onset), assuming *R=*2.6, the cases captured by surveillance must, if unisolated, be expected to cause 61% of onward transmission to achieve local elimination by surveillance and isolation alone (see Text S2).

## Discussion

This analysis of early SARS-CoV-2 cases and their close contacts in Shenzhen China, provides insights into the natural history, transmission and control of this disease. The values estimated provide the evidentiary foundation for predicting the impact of this virus, evaluating control measures, and guiding the global response. Analysis of how cases are detected, and use of data on individuals exposed but not infected, allow us to show that infection rates in young children are no lower than the population average (even if rates of clinical disease are). We are able to directly estimate critical transmission parameters, and show that, at least among observed contacts, transmission rates are low. Estimates of the distribution of time between symptom onset and case isolation by surveillance type reveal that heightened surveillance combined with case isolation could plausibly account for these low rates of transmission. These results paint a positive picture of the impact of heightened surveillance and isolation in Shenzhen. However, uncertainty in the number of asymptomatic cases missed by surveillance and their ability to transmit must temper any hopes of stopping the COVID-19 epidemic by this means.

This work further supports the picture of COVID-19 as a disease with a fairly short incubation period (mean 4-6 days) but a long clinical course ^2,7,19^, with patients taking many weeks to die or recover. It should be noted, however, that we estimate a higher proportion of cases taking 14 days or more to develop symptoms (9%) than previous studies ^6,7^.

Focusing on cases detected through contact-based surveillance adds nuance to previous characterizations of COVID-19. Since PCR testing of contacts is near universal, we can assume these cases are more reflective of the average SARS-CoV-2 infection than cases detected through symptomatic surveillance. In the contact-based surveillance group, any tendency for cases to be male or older (beyond the underlying population distribution, see Table S3) disappears. Further, in this group, 20% were asymptomatic at the time of first clinical assessment and nearly 30% did not have fever. This is consistent with a reasonably high rate of asymptomatic carriage, but less than suggested by some modeling studies^18^, though PCR has imperfect sensitivity^20^.

In Shenzhen, SARS-CoV-2 transmission is most likely between very close contacts, such as individuals sharing a household. However, even in this group less than 1 in 6 contacts were infected; and, overall, we observed far less than one (0.4) onward transmission per primary case. As noted above, low transmission levels may in part be due to the impact of isolation and surveillance; but it is equally likely unobserved transmission is playing some rule. We also estimate reasonably high rates of overdispersion in the number of cases each individual causes, leaving open the possibility that large COVID-19 clusters occur even if surveillance and isolation are forcing *R* below one; events that could potentially overwhelm the surveillance system.

This work has numerous limitations. As in any active outbreak response, the data were collected by multiple teams under protocols that, by necessity, changed as the situation developed. Hence, there may be noise and inconsistency in definitions. Of note, the definition of a confirmed case changed to require symptoms near the end of our analysis period (Feb. 7); but sensitivity analyses show that truncating the data at this point does not qualitatively impact results. It is, likewise, impossible to identify every potential contact an individual has, so contact tracing focuses on those close contacts most likely to be infected; hence our observed *R* is assuredly less than the true *R* in the population. Asymptomatic travellers will be missed by symptom-based surveillance; and, even if tested, some asymptomatic contacts may be missed due to the imperfect sensitivity of the PCR test ^20^.

As SARS-CoV-19 continues to spread, it is important that we continue to expand our knowledge about its transmission and natural history. Data from the early phase of local outbreaks, when detailed contact tracing is possible and sources of infection can still be reliably inferred, are particularly powerful for estimating critical values. This is especially true when information on uninfected contacts and mode of detection are used, as we have done here. The resulting estimates provide critical inputs for interpreting surveillance data, evaluating interventions, and setting public health policy.

## Data Availability

Linelist data contains PHI and cannot be made available. The authors are working to create deidentified summary data sets that will be made available when completed.

## ACKNOWLEDGEMENT

We thank Andrew Azman, Derek Cummings, Steve Lauer, Jacco Wallinga, and Michael Mina for advice and input on the manuscript and analyses. We thank all patients, close contacts, and their families involved in the study; as well as the front line medical staff and public health workers who collected this critical data.

## AUTHOR CONTRIBUTION

YW, SM, XZ, ZZ, XL, WG, LW, CC, XT, XW, YW, and SH collected the data JL, QB, SAT, and CY performed statistical analyses, and drafted the manuscript and figures TZ, BS, YS, JZ, TM, and CY cleaned the data QB, MT, JL, and TF conceived the study and supervised the collection of data

## SUPPLEMENTARY MATERIAL

**Table S1:** Demographic and clinical characteristics of cases by mode of case detection

| Symptom | Value | Contact-based<br>(N=87) | Symptom-based (N=292) | Unknown/<br>other (N=12) | Total (N=391) | P-value |
| --- | --- | --- | --- | --- | --- | --- |
| chill | no | 85 (97.7%) | 272 (93.2%) | 12 (100.0%) | 369 (94.4%) | 0.27 |
|  | yes | 2 (2.3%) | 20 (6.8%) | 0 (0.0%) | 22 (5.6%) |  |
| shortness of breath | no | 86 (98.9%) | 281 (96.2%) | 12 (100.0%) | 379 (96.9%) | 0.42 |
|  | yes | 1 (1.1%) | 11 (3.8%) | 0 (0.0%) | 12 (3.1%) |  |
| difficulty breathing | no | 87 (100.0%) | 285 (97.6%) | 11 (91.7%) | 383 (98.0%) | 0.35 |
|  | yes | 0 (0.0%) | 7 (2.4%) | 1 (8.3%) | 8 (2.0%) |  |
| chest tightness | no | 85 (97.7%) | 290 (99.3%) | 12 (100.0%) | 387 (99.0%) | 0.17 |
|  | yes | 2 (2.3%) | 2 (0.7%) | 0 (0.0%) | 4 (1.0%) |  |
| chest pain | no | 85 (97.7%) | 281 (96.2%) | 12 (100.0%) | 378 (96.7%) | 0.77 |
|  | yes | 2 (2.3%) | 11 (3.8%) | 0 (0.0%) | 13 (3.3%) |  |
| conjunctivitis | no | 87 (100.0%) | 290 (99.3%) | 12 (100.0%) | 389 (99.5%) | 1.00 |
|  | yes | 0 (0.0%) | 2 (0.7%) | 0 (0.0%) | 2 (0.5%) |  |
| nausea | no | 87 (100.0%) | 292 (100.0%) | 12 (100.0%) | 391 (100.0%) | 1.00 |
|  | yes | 0 (0.0%) | 0 (0.0%) | 0 (0.0%) | 0 (0.0%) |  |
| vomit | no | 87 (100.0%) | 289 (99.0%) | 12 (100.0%) | 388 (99.2%) | 0.64 |
|  | yes | 0 (0.0%) | 3 (1.0%) | 0 (0.0%) | 3 (0.8%) |  |
| diarrhea | no | 86 (98.9%) | 290 (99.3%) | 12 (100.0%) | 388 (99.2%) | 0.57 |
|  | yes | 1 (1.1%) | 2 (0.7%) | 0 (0.0%) | 3 (0.8%) |  |
| stomachache | no | 83 (95.4%) | 275 (94.2%) | 12 (100.0%) | 370 (94.6%) | 0.78 |
|  | yes | 4 (4.6%) | 17 (5.8%) | 0 (0.0%) | 21 (5.4%) |  |
| cough | no | 67 (77.0%) | 165 (56.5%) | 6 (50.0%) | 238 (60.9%) | 0.00 |
|  | yes | 20 (23.0%) | 127 (43.5%) | 6 (50.0%) | 153 (39.1%) |  |
| runny nose | no | 86 (98.9%) | 273 (93.5%) | 12 (100.0%) | 371 (94.9%) | 0.14 |
|  | yes | 1 (1.1%) | 19 (6.5%) | 0 (0.0%) | 20 (5.1%) |  |
| sore throat | no | 83 (95.4%) | 273 (93.5%) | 9 (75.0%) | 365 (93.4%) | 0.60 |
|  | yes | 4 (4.6%) | 19 (6.5%) | 3 (25.0%) | 26 (6.6%) |  |
| headache | no | 77 (88.5%) | 241 (82.5%) | 10 (83.3%) | 328 (83.9%) | 0.25 |
|  | yes | 10 (11.5%) | 51 (17.5%) | 2 (16.7%) | 63 (16.1%) |  |
| fatigue | no | 80 (92.0%) | 247 (84.6%) | 11 (91.7%) | 338 (86.4%) | 0.17 |
|  | yes | 7 (8.0%) | 45 (15.4%) | 1 (8.3%) | 53 (13.6%) |  |
| muscle soreness | no | 81 (93.1%) | 223 (76.4%) | 11 (91.7%) | 315 (80.6%) | 0.01 |
|  | yes | 6 (6.9%) | 69 (23.6%) | 1 (8.3%) | 76 (19.4%) |  |
| joint soreness | no | 83 (95.4%) | 247 (84.6%) | 12 (100.0%) | 342 (87.5%) | 0.01 |
|  | yes | 4 (4.6%) | 45 (15.4%) | 0 (0.0%) | 49 (12.5%) |  |

**Table S2:** Distributional fits to key COVID-19 distributions.

| Time lag | Distribution | Parameter 1 | Parameter 2 | Mean | 5% | 50% | 95% |
| --- | --- | --- | --- | --- | --- | --- | --- |
| Incubation period | lognormal | 1.57 (1.44,1.69) | 0.65 (0.56,0.73) | 5.95 (4.94,7.11) | 1.64 (1.33,2.04) | 4.80 (4.22,5.44) | 14.04 (12.19,15.90) |
| Serial interval | gamma | 2.29 (1.77,3.34) | 0.36 (0.26,0.57) | 6.29 (5.17,7.56) | 1.32 (0.92,1.87) | 5.41 (4.43,6.49) | 14.3 (11.12,17.57) |
| Onset to PCR confirmation;<br>among contact-based | gamma | 3.2 (2.43,4.76) | 1 (0.71,1.57) | 3.18 (2.65,3.76) | 0.92 (0.7,1.26) | 2.86 (2.4,3.37) | 6.56 (5.25,8.01) |
| Onset to PCR confirmation;<br>among symptom-based | gamma | 2.12 (1.87,2.45) | 0.39 (0.33,0.46) | 5.46 (4.99,5.92) | 1.04 (0.88,1.26) | 4.63 (4.23,5.03) | 12.71 (11.51,13.82) |
| Onset to hospitalization;<br>among contact-based | lognormal | 0.74 (0.55,0.95) | 0.64 (0.55,0.71) | 2.57 (2.06,3.16) | 0.73 (0.61,0.93) | 2.09 (1.73,2.58) | 6.03 (4.49,7.53) |
| Onset to hospitalization;<br>among symptom-based | lognormal | 1.23 (1.12,1.33) | 0.79 (0.74,0.83) | 4.64 (4.13,5.1) | 0.93 (0.82,1.08) | 3.41 (3.06,3.78) | 12.42 (10.89,13.77) |
| Onset to isolation;<br>among contact-based | lognormal | 0.77 (0.53,0.97) | 0.67 (0.56,0.75) | 2.71 (2.08,3.31) | 0.72 (0.58,0.96) | 2.17 (1.71,2.64) | 6.52 (4.69,8.24) |
| Onset to isolation;<br>among symptom-based | lognormal | 1.22 (1.12,1.31) | 0.78 (0.73,0.83) | 4.58 (4.13,5.02) | 0.94 (0.82,1.08) | 3.38 (3.07,3.69) | 12.19 (10.79,13.62) |
| Arrival to symptom onset;<br>among onset after arrival | lognormal | 1.22 (1.1,1.34) | 0.85 (0.79,0.91) | 4.87 (4.24,5.49) | 0.83 (0.71,0.97) | 3.38 (2.99,3.81) | 13.79 (11.75,15.86) |
| Arrival to confirmation;<br>among onset on or before<br>arrival | weibull | 1.28 (1.04,1.59) | 4.8 (4.05,5.58) | 4.5 (3.81,5.07) | 0.47 (0.25,0.83) | 3.61 (2.92,4.35) | 11.32 (9.56,13.06) |
| Arrival to isolation;<br>among onset on or before<br>arrival | gamma | 0.39 (0.3,0.54) | 0.13 (0.1,0.18) | 3.05 (2.49,3.67) | 0 (0,0.02) | 1.09 (0.67,1.66) | 12.75 (10.57,14.84) |

**Table S3:** Comparison of age distribution of cases with Shenzhen 2010 census (source http://www.stats.gov.cn/english/)

| Age category | N | Proportion | Contact-based | Symptom-based |
| --- | --- | --- | --- | --- |
| 0-9 years | 736,978 | 7% | 13 (14.9%) | 6 (2.1%) |
| 10-19 years | 1,058,098 | 10% | 5 (5.7%) | 6 (2.1%) |
| 20-29 years | 3,783,127 | 37% | 11 (12.6%) | 23 (7.9%) |
| 30-39 years | 2,528,979 | 24% | 15 (17.2%) | 71 (24.3%) |
| 40-49 years | 1,478,974 | 14% | 9 (10.3%) | 49 (16.8%) |
| 50-59 years | 466,403 | 5% | 10 (11.5%) | 63 (21.6%) |
| 60-69 years | 192,595 | 2% | 20 (23.0%) | 60 (20.5%) |
| 70+ years | 113,227 | 1% | 4 (4.6%) | 14 (4.8%) |
| Total | 10,358,381 | 100% | 87 (100.0%) | 292 (100.0%) |

**Table S4:** Comparison of observed serial intervals by time between symptom onset and isolation.

| Time to isolation | Mean serial interval (95% CI) |
| --- | --- |
| 0-2 days | 3.6 (3.0, 4.2) |
| 3-5 days | 8.1 (5.3, 11.0) |
| 6 or more days | 8.0 (6.2, 9.7) |

**Table S5.** Time to recovery from symptom onset in days.

| Variable | Value | Time to recovery | 2.5% | 97.5% |
| --- | --- | --- | --- | --- |
| sex | female | 20.3 | 19.4 | 21.3 |
|  | male | 21.2 | 20.2 | 22.3 |
| age | 0-9 | 17.5 | 15.3 | 20.0 |
|  | 10-19 | 19.1 | 15.8 | 22.9 |
|  | 20-29 | 19.2 | 17.5 | 21.0 |
|  | 30-39 | 19.2 | 18.0 | 20.5 |
|  | 40-49 | 21.6 | 20.0 | 23.4 |
|  | 50-59 | 22.4 | 20.8 | 24.1 |
|  | 60-69 | 22.9 | 21.2 | 24.7 |
|  | 70+ | 22.5 | 19.1 | 26.3 |
| severity | mild | 20.1 | 19.0 | 21.3 |
|  | moderate | 20.3 | 19.5 | 21.1 |
|  | severe | 28.3 | 25.3 | 31.6 |
| mode of detection | contact-based | 19.3 | 17.9 | 20.9 |
|  | symptom-based | 21.2 | 20.4 | 22.0 |
| Total | Total | 20.8 | 20.1 | 21.5 |

**Figure S1:**
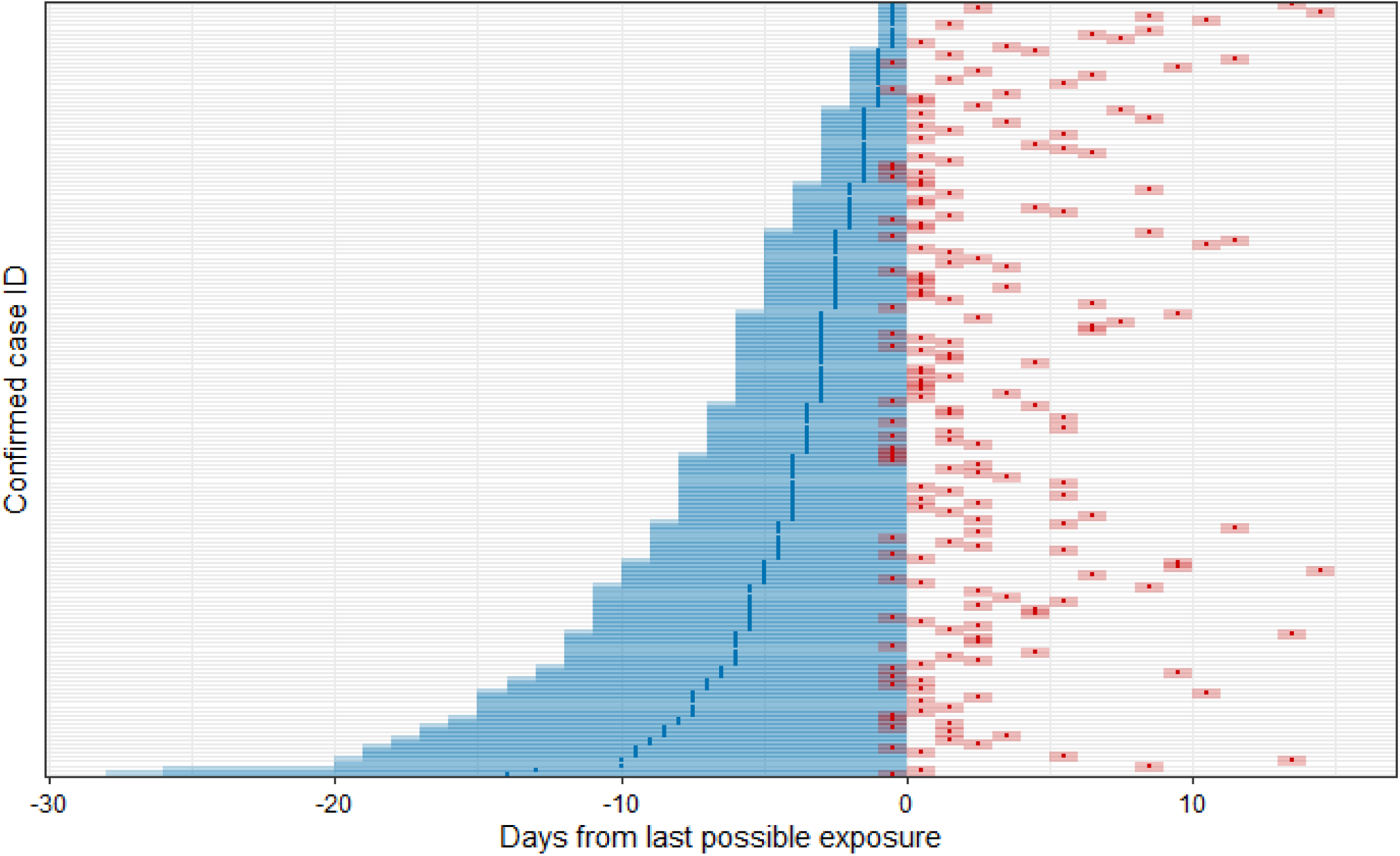
**The exposure and symptom onset windows 339 confirmed cases from Shenzhen, China**. Shaded regions represent the full possible interval of exposure (blue) and of symptom onset (red); points represent the midpoint of these intervals. The exposure and symptom onset windows are aligned relative to the right-bound of the exposure window.

**Figure S2:**
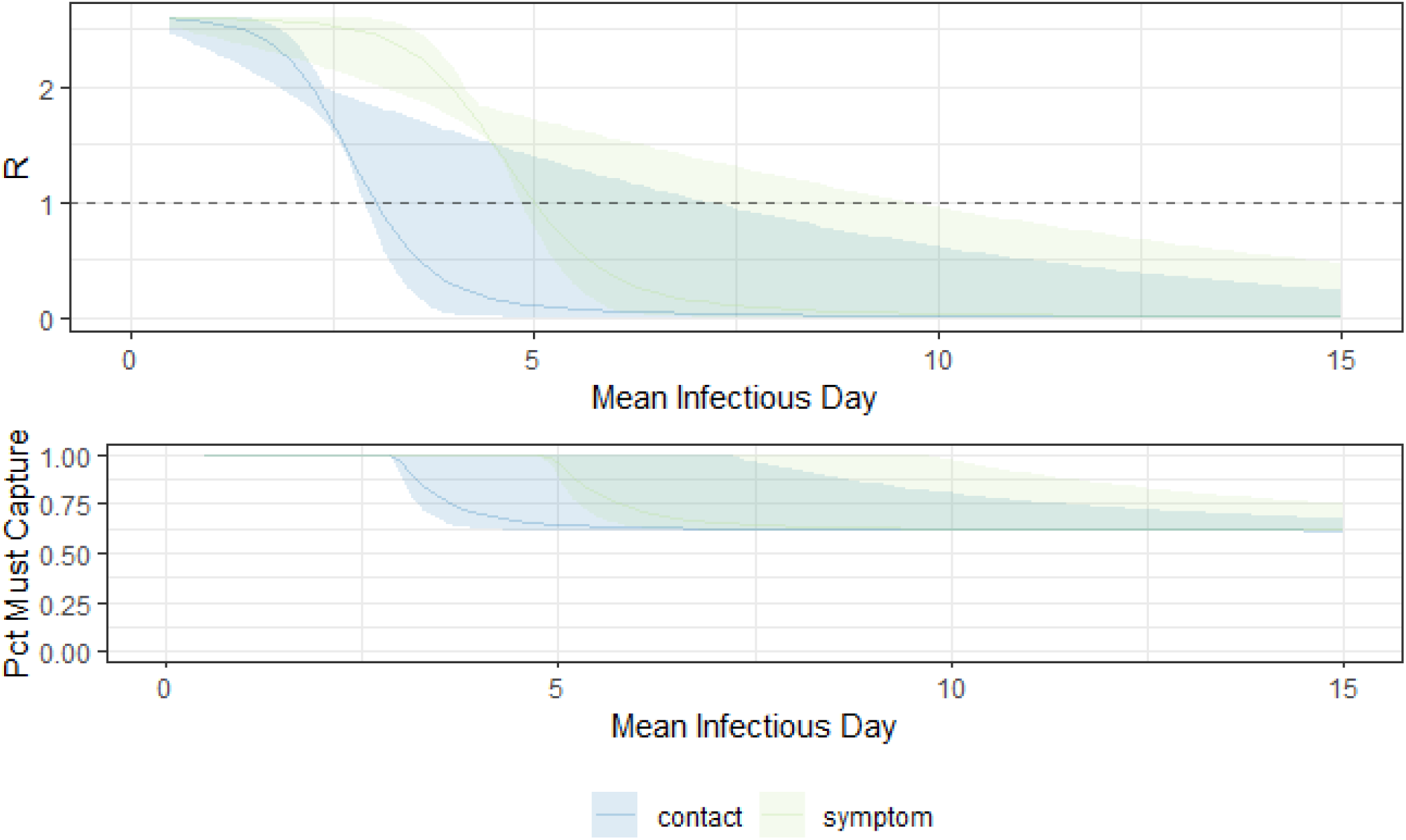
Effective *R* among those captured by surveillance (top) and proportion needed to be captured by surveillance to drive *R* less than one (bottom) by the weighted mean day of the infectious period. Weighting is by relative infectiousness, which is assumed to follow a gamma distribution. The shaded area covers all gamma distributions with a mean of that day and a rate parameter in the range of 0.1-10.

**Figure S3:**
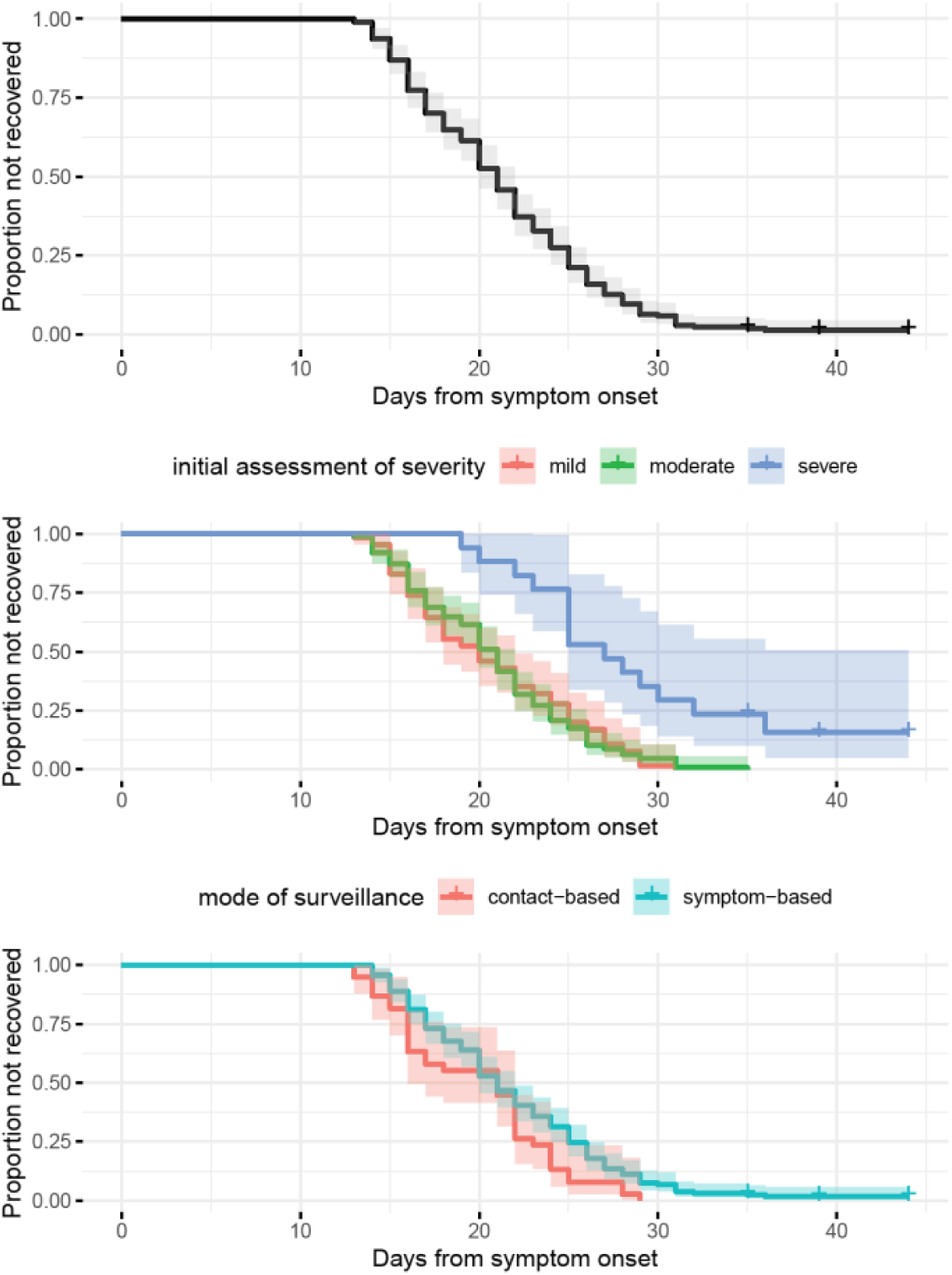
Time from symptom onset to recovery for all cases (top), by initial assessment of clinical severity (middle), and by mode of surveillance (bottom). Time from symptom onset to death was marked by “+” for the three patients who have died.

### Text S1: Data Extraction and Confirmation Details

By categorizing COVID-19 as a notifiable disease Class B, Chinese Law on the Prevention and Treatment of Infectious Diseases required all cases to be immediately reported to China’s Infectious Disease Information System. Each case was recorded into the system by local epidemiologists and public health professionals who did the field investigation and collected possible exposure related information. All data on COVID-19 case reported in Shenzhen were extracted from the Infectious Disease Information System by the end of February 12, 2020. Then personal information including demographics, symptoms, clinical outcome and severity and so on, were stripped to construct an anonymous dataset. All cases were included without sampling and no eligibility criteria were needed.

All laboratory confirmation of SARS-CoV-2 were done by Guangdong Center for Disease Prevention and Control (CDC) before Jan 30, 2020, and then only need to be done by Shenzhen CDC, when it obtained the qualification of laboratory-confirmation of 2019-nCoV from the authority. The RT-PCR assay was conducted in the BSL-2 laboratory of Shenzhen CDC, using the protocol established by the World Health Organization and China CDC.

### Text S2: Supplemental Calculation

Let *R*_0_ be the basic reproductive number, *ρ* be the percent of transmission due to cases potentially reachable by the surveillance system, and *γ* be the relative effective infectious period of those captured by surveillance. Then:

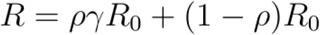

When *R* is below one, sustained outbreaks are impossible. Hence, for a known *R*_0_ and *γ* such that *γ R*_0_ < 1, we can calculate the proportion of transmission that must be from people who can be captured by surveillance as:

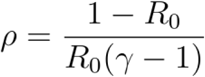

Assuming an *R*_0_ of 2.6 and that surveillance reduces *R*_0_ by a factor of 0.18, we find control is possible if 75% of people can be captured by surveillance.

